# On dynamics of fractional incommensurate model of Covid-19 with nonlinear saturated incidence rate

**DOI:** 10.1101/2021.07.18.21260711

**Authors:** Abdelouahed Alla hamou, Elhoussine Azroul, Zakia Hammouch, Abdelilah Lamrani alaoui

**Affiliations:** Laboratory of Mathematical Analysis and Applications, Faculty of Sciences Dhar Al Mahraz, B.P. 1796, 30000, Fez, Morocco; Ton Duc Thang University, Ho Chi Minh City, Vietnam; Faculty of Mathematics and education sciences, Harran University Sanliurfa, Turkey; Department of Mathematics, Regional Center of Education and Professional Training,B.P. 49, 30000 Fez, Morocco

**Keywords:** Atangana–Baleanu derivative, COVID-19, SQIR epidemic model, Fractional differential equations

## Abstract

In December 2019, a new virus belonging to the coronavirus strain has been discovered in Wuhan, China, this virus has attracted world-wide attention and it spread rapidly in the world, reaching nearly 216 countries in the world in November 2020. In this chapter, we study the fractional incommensurate SIQR (susceptible, infections,quarantined and removed) COVID-19 model with nonlinear saturated incidence rate using Atangana–Baleanu fractional derivatives. The existence and uniqueness of the solutions for the fractional model is proved using fixed point theorem, the model are shown to have two equilibrium point (disease-free and an endemic equilibrium). Some numerical simulations using Euler method are also carried out to support our theoretical results. We estimated the value of the fractional orders and the parameters of the proposed model using the least squares method.. Further, the sensitivity analysis of the parameter is performed as a result, our incommensurate model gives a good approximation to real data of COVID-19.

## 1. Introduction

At the end of December 2019, an unidentified virus was found in Hubei province, China. The responsible virus was later confirmed as a new coronavirus [1]. The World Health Organization (WHO) named the virus as the 2019 novel coronavirus (2019-nCoV) or COVID-19 or 11 February 2020. The first confirmed case of the virus was discovered on 17 November 2019 in Hubei. As of November 3, 2020, more than 47.58 million cases have been reported in 216 countries in the world, which resulted in more than 1,215,180 deaths. Over 34.185 million people recovered [2].

A fractional differential equations are equations where the order of the derivative is a real or complex number. Its first appearance is in a letter written to Guillaume de l’Hôpital by Gottfried Wilhelm Leibniz in 1695. There are several types of fractional derivatives in the literature. We mention them Riemann–Liouville derivative, Caputo derivative, Riesz derivative, Grünwald–Letnikov derivative, Hadamard derivative, Marchaud derivative, Weyl derivative and recently, other types of fractional derivative have appeared, the most important of which are Caputo-Fabrizio derivative and Atangana-Baleanu derivative, the latter has aroused the interest of many researchers in various fields, due to its efficiency in modeling real problems in several areas, namely epidemiological problems. In recent years researchers have been interested in studying some real problems in various fields using fractional calculus, such as physical problems [1, 3, 4], engineering mechanics [5], epidemiological models [6, 7,8,9,10], image processing [11, 12], chaos theory [13, 14] and others.

After the emergence of the Corona pandemic, researchers in various fields tried to study the evolution of this virus, how it is transmitted and provide the appropriate mathematical model for it. Many epidemiological models with fractional (non-integer order) derivative have been developed to understand the transmission of 2019-nCoV and other infectious diseases outbreak from various aspects. We mention them, Singh et al. [15] have been studied the fractional epidemiological model for computer viruses using CF-fractional derivative. The comparison between the classical and fractional SEIR epidemic model of Ebola virus was studied by Area et al. [7]. Tulu et al. [16] developed the Caputo fractional mathematical model of the Ebola virus with a quarantine strategy and the presence of the vaccine by using MCMC method. Dokuyucu and Dutta [8] examined the new Ebola virus model with CF-fractional operators. Kumar et al. [17] developed Caputo–Fabrizio fractional SIRS-SI model for the malaria disease. Singh et al. [6] studied the diabetes epidemic model using the operators proposed by Caputo and Fabrizio. Baleanu et al. [10] studied a time fractional SEIR model for COVID-19 with Caputo-Fabrizio derivative and an auxiliary parameter by evoking asymptomatic infected people and the COVID-19 case in the reservoir. Abdo et al. [18] used Atangana-Baleanu-Caputo operator to study the mathematical model of COVID-19. Thabet et al. [19] suggested a fractional SEIHDR model for COVID-19 using ABC fractional derivative. Zhang et al. [20] applied the fractional SEIRD Model to the real data of the 2019-nCoV for China using Caputo fractional derivative. ud Din et al. [9] studied a general SEIARW fractional model for the transmission of COVID-19 by using the new fractional operators of Caputo and Fabrizio. Higazy [21] suggested a fractional SIDARTHE epidemic model for 2019-nCoV by using CF-fractional operators. Some other outstanding studies of 2019-nCoV by fractional derivative have been made in [22, 23, 24, 25].

Always when a new virus appears, researchers look for effective solutions and methods to control the new virus, including vaccination, isolation, and quarantine. In the absence of the vaccine, the isolation and quarantine strategies remain effective to mitigate and eliminate the impact of the virus (see [26,27, 28,29,30,31,32,33]).

In [34] the authors suggested changing the fractional SIAR model for HIV/AIDS with treatment compartment from classical commensurate model to incommensurate Caputo-Fabrizio fractional model by giving each equation a different order from the other equations. In this chapter we will study the general SQIR epidemiological model for COVID-19 with saturated incidence rate and using ABC-fractional operator. We will compare the results obtained for the fractional model with the classical model by predicting the model using Least Squares method and real data of Mexico from April 12 to October 28, 2020. Then the novelty of this paper is to use a different fractional order for each equation of the SQIR epidemic system with AB-fractional operators, this is fractional incommensurate SQIR model with time fractional derivative and saturated incidence rate.

The paper is organized as follows. In section 2, we present the Background of Atangana-Baleanu derivative and integral operators. The classical and fractional models are formulated in Section 3. The fixed point theorem is applied to prove existence and uniqueness results in section 4. The disease-free equilibrium and their local stability are given in section 5. In section 6 the Euler numerical method is used to determine the numerical solutions of the fractional proposed model. The model parameters are estimated according to real data of Mexico and the influence of fractional order has been shown in section 7. Finally, we gave some remarks and conclusions in section 8.

## 2. Background of Atangana-Baleanu operators

We first give some preliminary on the definitions of fractional derivatives will be used in this paper. [35].

### Definition 1

*Let* (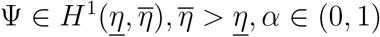), *then the Atangana–Baleanu-Caputo (ABC for short) derivative is given as*

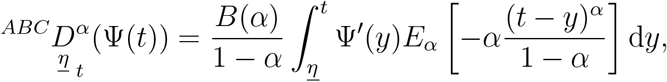

*where B is a function fulfills the condition B*(1) = *B*(0) = 1. *and E*_*α*_ *is the generalized Mittag–Leffler function given by*

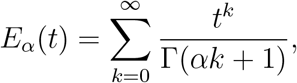

*where* Γ *is Gamma function*.

The corresponding integral is given by the flowing definition.

### Definition 2

*The AB-fractional integral associated to the Atangana-Baleanu fractional derivative is defined as*

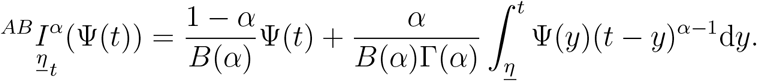

### Remark 1

When *α* → 0 in the above definition we find the initial function. Moreover, we find the Riemann integral if the fractional order *α* turns to 1.

## 3. The SQIR model formulation

We consider in this part the model of COVID-19. The population is divided into four compartments, the first is the susceptible class *S*(*t*), Infected class *I*(*t*), Quarantined class *Q*(*t*) and Recovered class *R*(*t*).

The transfer diagram for this model is described by Figure 1 and the classical version of this model formulated by the flowing system of four nonlinear ODEs :

**Figure 1:**
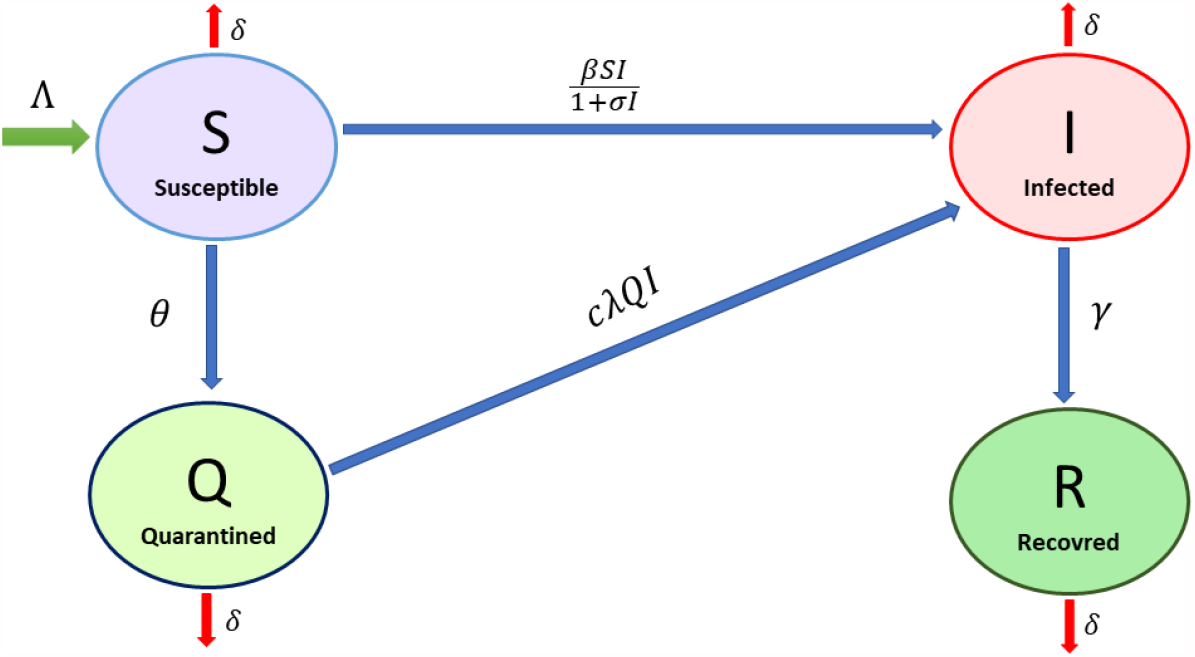
Schematic diagram of 2019-nCoV model and parameters.

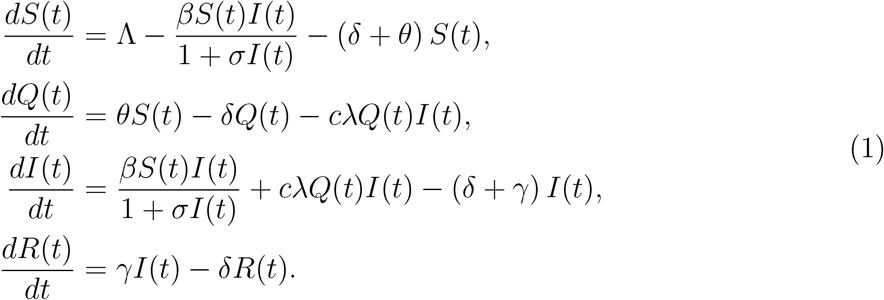

with the initial conditions *S*(0) = *S*_0_ ⩾ 0, *Q*(0) = *Q*_0_ ⩾ 0, *I*(0) = *I*_0_ ⩾ 0 and *R*(0) = *R*_0_ ⩾ 0.

### 3.1. Fractional model

In this section, we change the classical derivative in (1) by ABC-fractional derivatives mentioned in Section 2.

Now remplacing the classical derivative in (1) by Atangana-Baleanu-Caputo derivative we obtain by moment closure, the following system of ABC-fractional ODEs

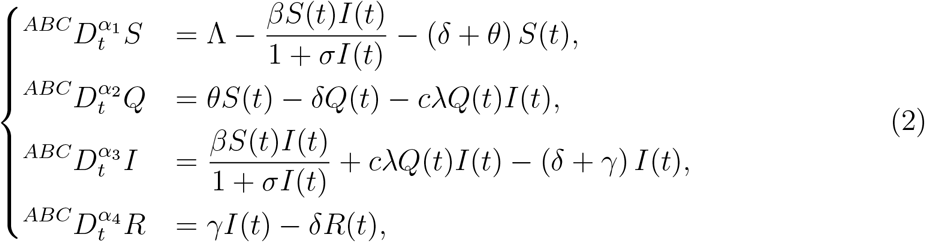

with the initial conditions *S*(0) = *S*_0_ ⩾ 0, *Q*(0) = *Q*_0_ ⩾ 0, *I*(0) = *I*_0_ ⩾ 0, *R*(0) = *R*_0_ ⩾ 0 and 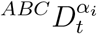, *i* = 1, 2, 3, 4, is the ABC-fractional operator of order 0 *< α*_*i*_ *<* 1.

The total population *N* (*t*) = *S*(*t*) + *I*(*t*) + *R*(*t*) + *Q*(*t*). Then

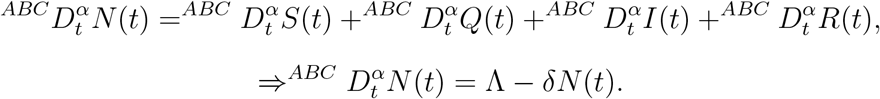

In the absence of the disease, 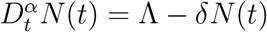, this implies

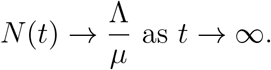

The dynamics of the fractional COVID-19 model will be studied in a biologically feasible set denoted by

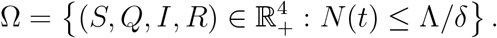

## 4. Existence and uniqueness of solutions of the ABC-fractional model

In this section, we prove the existence of the system of solutions by applying the fixedpoint theorem.

Let ℋ = (*C*([0, *T*]))^4^, and *C*([0, *T*]) be a Banach space of all real-valued, continuous functions on the interval [0, *T*] with the norm

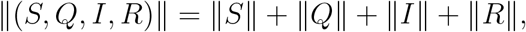

where ‖ · ‖ denote the supremum norm in *C*([0, *T*]).

We use the following notations for the right side of the COVID-19 models

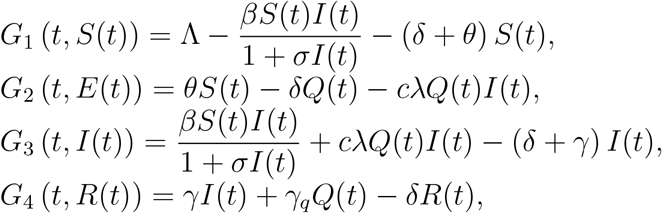

by applying the definition of fractional integral to the Equation (1) and using the properties of fractional calculus we get

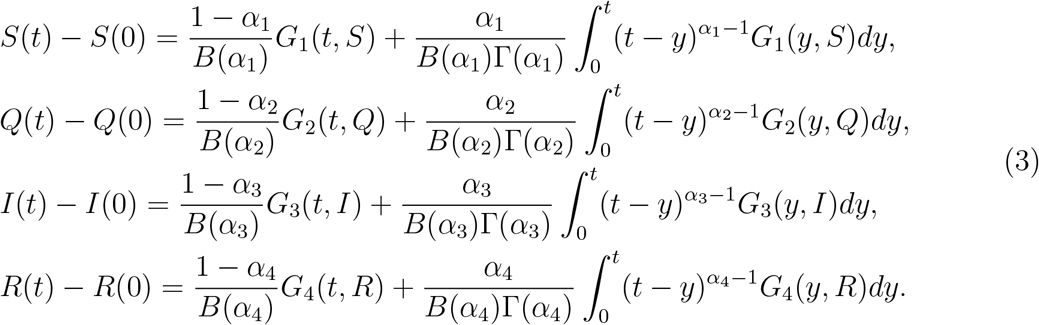

To prove the following theorems, we will assume that ‖*S*(*t*) ‖ ≤ *C*_1_, ‖*Q*(*t*) ‖ ≤ *C*_2_, *‖I*(*t*) ‖ ≤ *C*_3_ and *‖R*(*t*) *‖* ≤ *C*_4_, where *C*_*i*_, *i* = 1, …, 4, are some positive constants. Denote

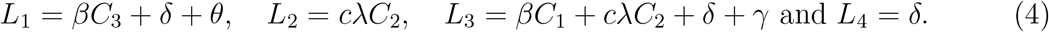

### Theorem 3

*Assume that the following inequality is verified*

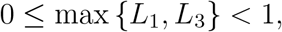

*then the kernels G*_1_, *G*_2_, *G*_3_ *and G*_4_ *fulfill Lipschitz conditions, furthermore are contraction mappings*.

Proof. Let *S*_1_ and *S*_2_ be two functions of susceptible idividuals, then we have

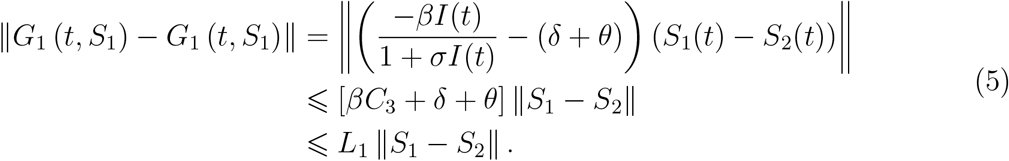

where *L*_1_ is defined in (4). Similar results for the kernels *G*_2_, *G*_3_, *G*_4_, and *G*_5_ can be obtained using {*Q*_1_, *Q*_2_}, {*I*_1_, *I*_2_} and {*R*_1_, *R*_2_} respectively, as follows

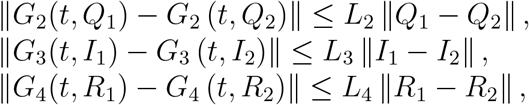

where *L*_2_, *L*_3_ and *L*_4_ are expressed in (4). Therefore, the Lipschitz conditions are fulfilled for *G*_2_, *G*_3_, and *G*_4_. In addition, since 0 ≤ max{*L*_1_, *L*_2_, *L*_3_, *L*_4_} *<* 1, the kernels are contractions.

### Theorem 4

*Assume that the following inequality fulfill:*

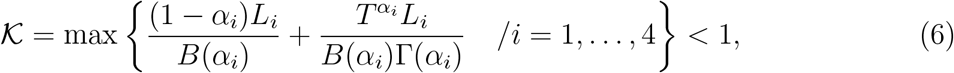

*then the COVID-19 model with ABC-fractional operator (2) has a unique solution*.

Proof. The proof of this theorem is based on the fixed point method and using Picard operator. We consider the following notations

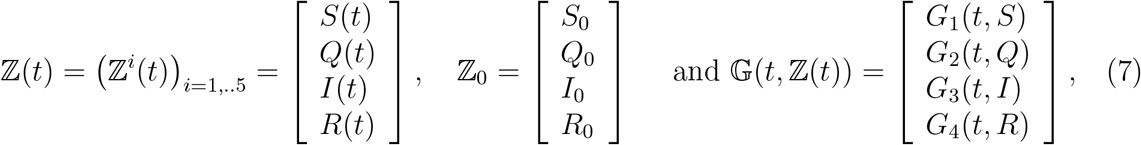

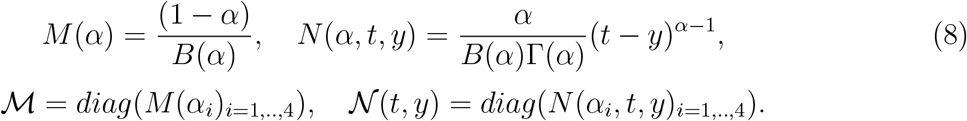

Keeping in mind the above notation, system (2) may be expressed as

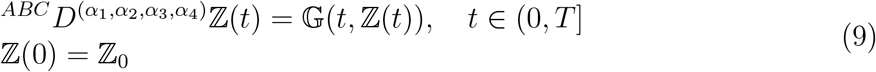

where

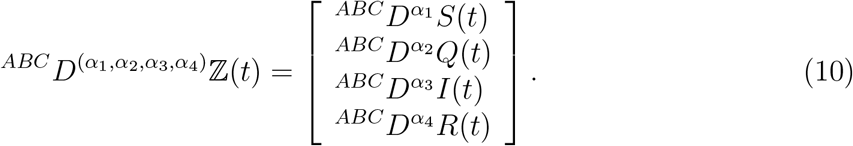

Now using (3) and according to the above notation we get the following fractional integral equation

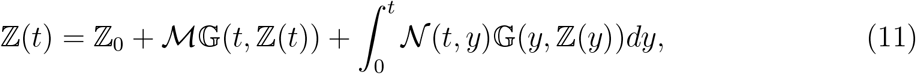

define

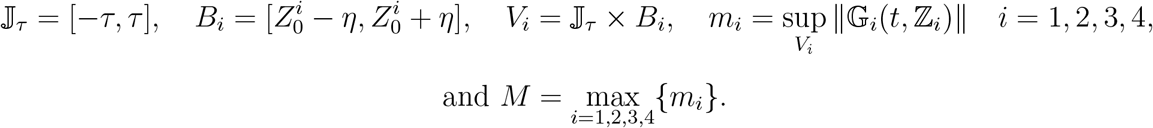

Now using the metric on C (𝕁, *B*_1_, *B*_2_, *B*_3_, *B*_4_) induced by the norm given as

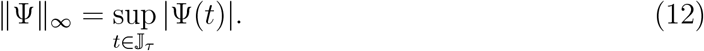

We define Picard’s operator as

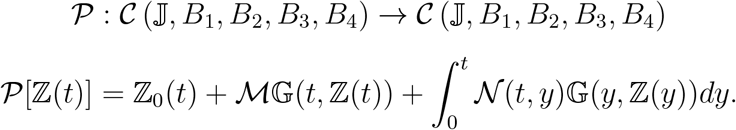

Now let’s try to prove that 𝒫 is a contraction.

Let 𝕫_1_, *𝕫*_2_ ∈ C (𝕁, *B*_1_, *B*_2_, *B*_3_, *B*_4_) two functions, then we have

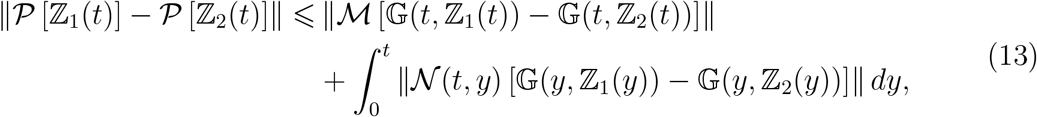

by using the triangular inequality and the Lipschitz condition we will find

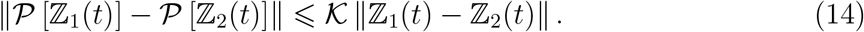

If condition (6) fulfill then 𝒫 is a contraction on (𝕁, *B*_1_, *B*_2_, *B*_3_, *B*_4_). Then by Banach fixed point theorem 𝒫 has a unique fixed point. Hence, the fractional COVID-19 model (2) has a unique continuous solution.

## 5. Basic reproduction number, existence and stability of equilibria

In this section we determine the equilibria of the COVID-19 models, for that we solve :

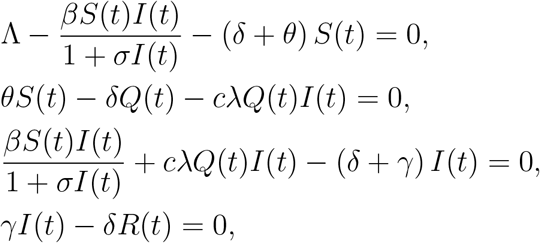

then the disease-free equilibrium is given by

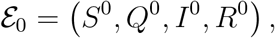

where 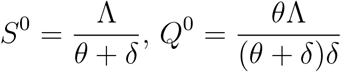 and *I*^0^ = *R*^0^ = 0.

Now let’s determine the expression of basic reproduction number denoted by ℛ_0_, by using the method presented in [36], we get

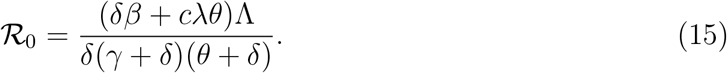

Furthermore, the system (2) has a unique endemic steady state

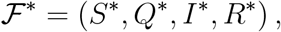

such as

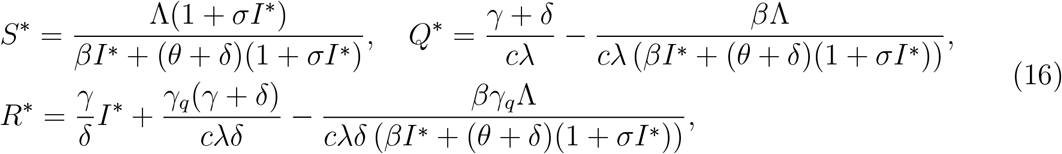

where the value of *I*\* is the positive solution of the following algebraic equation

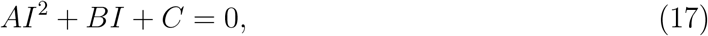

with

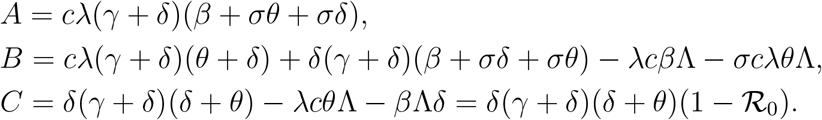

It remains to show that the solution of (17) is real and positive. We note

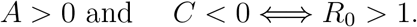

Let the discriminant of (17) be Δ, so that Δ = *B*^2^ −4*AC*.We conclude the following results for the endemic equilibrium.

### Theorem 5

*The system (2) has a unique endemic steady state whenever* ℛ_0_ *>* 1 *given by*

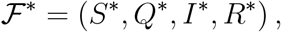

*where the expressions of S*\*, *Q*\**and R*\* *are given in (16) and* 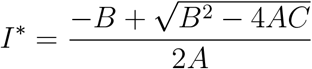.

### 5.1. Local stability analysis for the fractional model

In this part we prove the local asymptotic stability of the equilibria of the commensurate model of (2), that is to say in the case where the equations have the same fractional order *α* ∈ (0, 1). Since *R*(*t*) is not present in first three equations, then we can write the system (2) as

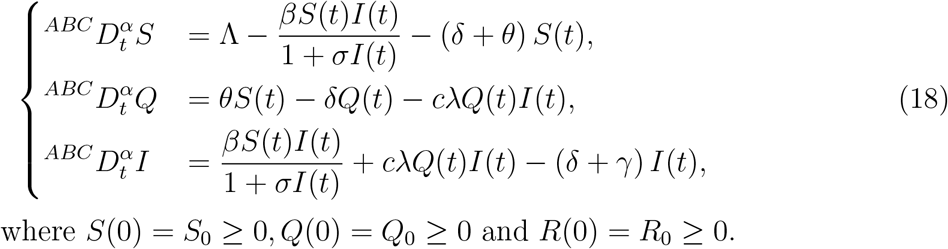

To show the local asymptotic stability of Atangana-Baleanu-Caputo model, it suffices to show that all the eigenvalues *λ*_1_, *λ*_2_ and *λ*_3_ of the jacobian matrix evaluated at the equilibrium point satisfy the condition

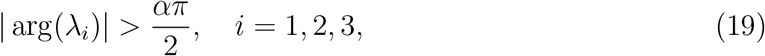

this result is mentioned in [37, Lemma 3.1.] and other references [38, 39].

#### Theorem 6

*If* ℛ_0_ *<* 1, *then ℱ*_0_ = (*S*^0^, *Q*^0^, *I*^0^) *is locally asymptotically and if ℛ*_0_ *>* 1, *then* F* = (*S*\*, *E*\*, *I*\*) *is locally asymptotically stable*.

Proof. The Jacobian matrix *J* (*ℱ*_0_) for the fractional model given by (18), at the equilibrium *ℱ*_0_ is given by:

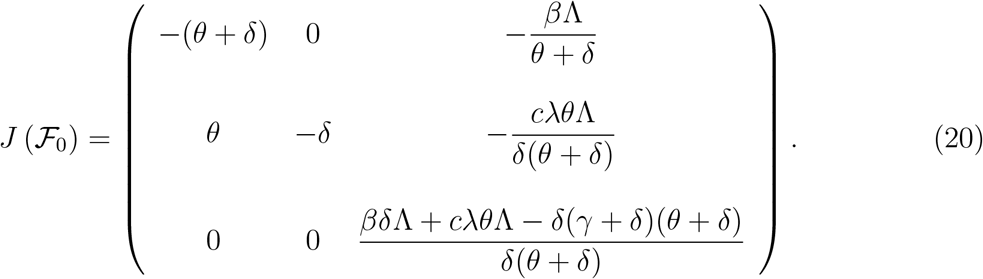

We find the eigenvalues by solving the following characteristic equation

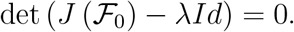

We obtain the equation

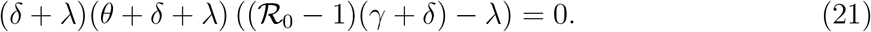

The eigenvalues of matrix *J* (*ℱ*_0_) are

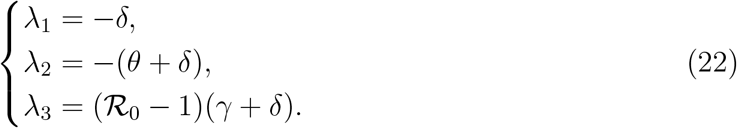

If *ℛ*_0_ *<* 1 then *λ*_1_, *λ*_2_ and *λ*_3_ are real negative roots. Then

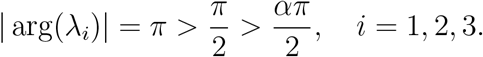

Hence the disease free equilibrium of the fractional model (18) is locally asymptotically stable.

Now, we prove the local stability of the endemic equilibrium point. We consider the Jacobian matrix *J* (*ℱ*\*) for the fractional COVID-19 model given by (18), at the equilibrium *ℱ*\* is given by

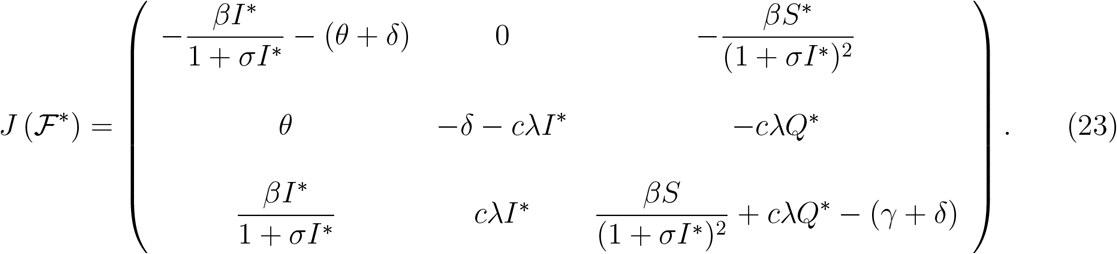

The eigenvalues of this matrix can be obtained by solving the flowing characteristic equation

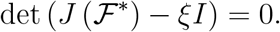

Therefore, we find the following characteristic equation:

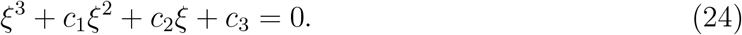

where

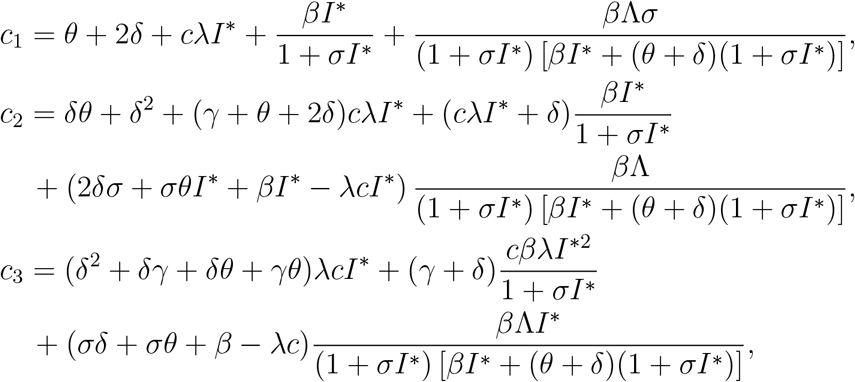

Let us denote by

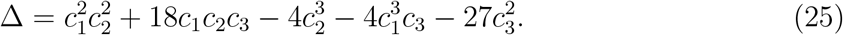

its discriminant, the following lemma completes the proof of theorem.

#### Lemma 7

*Asume that* ℛ_0_ *>* 1 *and one of the following conditions is satisfied:*

1. Δ *>* 0, *c*_1_ *>* 0, *c*_2_ *>* 0 *and c*_1_*c*_2_ − *c*_3_ *>* 0.
2. 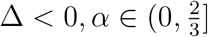, *c*_1_ ≥ 0, *c*_2_ ≥ 0 *and c*_3_ *>* 0.

*Then the equilibrium ℱ*\* *of the fractional COVID-19 model with ABC derivative is locally asymptotically stable*

Proof. The detailed proof of this lemma is similar to [40, Proposition 1.].

## 6. Numerical method

To develop approximation schemes for the fractional model, we will use the Fractional version of Euler Method [41]. To applied this method, using same notation of section 4 and we consider the incommensurate system of ABC-fractional differential equations

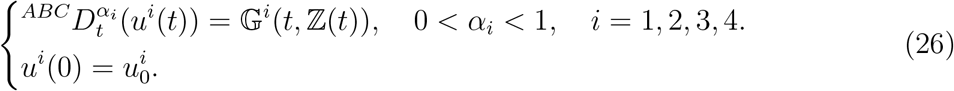

We consider *t*_0_ = 0 *< t*_1_ *<* … *t*_*k*_ … *< t*_*N*_ = *T* an uniform time discretization of an interval [0, *T*] in *N* + 1 points. Let 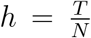 is the time step size. We denote 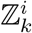 as the numerical approximation of ℤ ^*i*^ (*t*_*k*_).

Using AB-fractional integral, we have

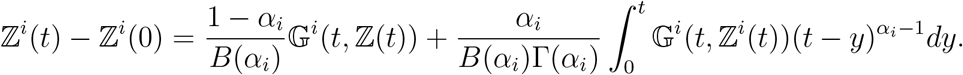

This implies

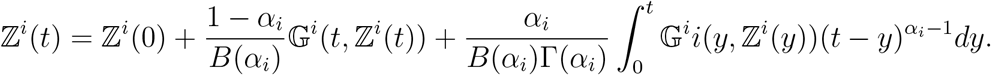

At the point *t*_*n*+1_ = (*n* + 1)*h*, where *h* = *t*_*n*+1_ − *t*_*n*_, we have

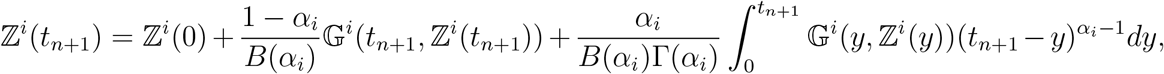

after using the fractional Euler method [41], we obtain

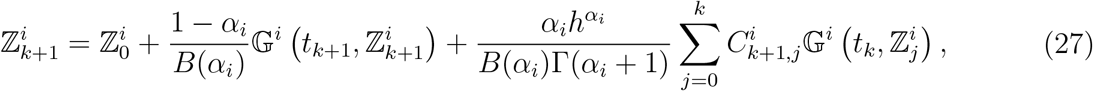

where the coefficients 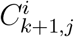, *j* = 0,…,*k*, are given by

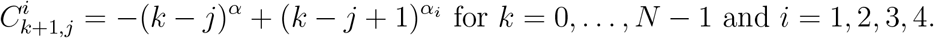

According to [41], the numerical scheme (27) for the fractional model (2) is conditionally stable. Now we will give a theorem for estimating the error between exact and numerical solution of the proposed model.

≤ {}

### Theorem 8

*Let ℤ*(*t*) *and* (ℤ_*k*_)_*k*_ *be respectively the exact and approximate solutions of the fractional incommensurate model (2). If* 0 ≤ max {*L*_1_, *L*_3_} *<* 1, *where L*_1_ *and L*_3_ *are given in (4), then we have*

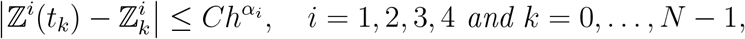

*where C is an independent positive constant free of k and h*.

Proof. The proof is a direct conclusion of [41, Theorem 4].

## 7. Numerical simulation and calibration of the fractional model

### 7.1. Estimation of model parameters and best fit of fractional orders

The model calibration problem seeks to estimate the model parameters which, to some extent, to make the error between the observed values and the predicted values as small as possible, we use for that the Least Squares method to produce the values of the parameters and the fractional multi-order. Some model parameters can be known by these real values, on the other hand some parameters are unknown and need to be estimated, the same for the initial conditions of the COVID-19 model.

We consider the model solution *Z*(*t*) = (*S*(*t*), *Q*(*t*), *I*(*t*), *R*(*t*)), depending on the vector of parameters *ξ* = (*β, c, λ, θ, δ, γ, σ*, Λ), the initial conditions *Z*_0_ = (*S*_0_, *Q*_0_, *I*_0_, *R*_0_) and the fractional orders *α* = (*α*_1_, *α*_2_, *α*_3_, *α*_4_, *α*_5_). The initial conditions *S*_0_, *I*_0_ and *R*_0_ are known from data of COVID-19, but *Q*_0_ is unknown so it is necessary to estimate its value.

The vector of parameters belongs to a set given by

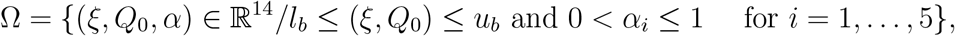

where *l*_*b*_ and *u*_*b*_ are of lower and upper vector values for the model parameters respectively.

Let the vector Θ ∈ ℝ^*N*^ of the observation data at given times *t*_*k*_, *k* = 1, … *N* and a prediction vector Ψ(*ξ, Q*_0_, *α*), the calibration aims at finding a vector 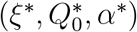 such that

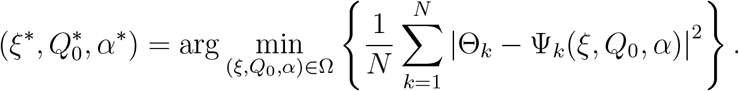

The aim is to minimize the objective function

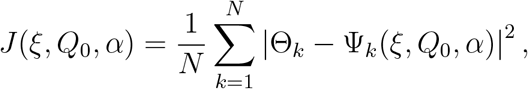

subject to (*ξ, Q*_0_, *α*) ∈ Θ and Equations (2) to obtain the best estimate of parameters and the vector of fractional orders α.

Now we consider the real data of COVID-19 for Mexico (that are found on the WHO web-page) to estimate the model parameters using the least squares method. In the first time we estimate the parameters for the classical model (1), second for the commensurate model which the four equations of the fractional model have the same order and finally the incommensurate model (2) when each equation has a different order.

The resulting fitted curves for reported infected and recovered cases of COVID-19 in Mexico is shown in Figure 2 for the classical model, in Figure 3 for the commensurate model and in Figure 4 for the incommensurate model, where the best fitted values of the parameters are depicted in Table 3 and the estimated values of the fractional orders of the two fractional models are given in Table 4. Furthermore, the initial conditions are given in the table 2.

**Table 1:** Parameters of the model.

| Parameters | Description |
| --- | --- |
| S(t) | Susceptible class |
| I(t) | Infected class |
| Q(t) | Quarantine class |
| R(t) | Recovered class |
| c | Contact rate |
| $\delta$ | Natural death rate |
| $\sigma$ | the constant of saturation |
| $\Lambda$ | Recruitment rate |
| $\beta$ | incidence rate |
| $\gamma$ | Recovery rate from COVID-19 |
| $\lambda$ | The interaction between S and Q |
| $\theta$ | Quarantine rate of susceptible |

**Table 2:** Estimated fractional orders.

| Model/Initial values | $N_0$ [2] | $S_0$ (Calculated) | $I_0$ [2] | $R_0$ [2] | $Q_0$ (Estimated) |
| --- | --- | --- | --- | --- | --- |
| Classical | 129387968 | 125670303 | 2643 | 1722 | 3713300 |
| Commensurate | 129387968 | 120741203 | 2643 | 1722 | 8642400 |
| Incommensurate | 129387968 | 37805603 | 2643 | 1722 | 91578000 |

**Table 3:** Estimated model parameters.

| Parameters | Classical model | Commensurate model | Incommensurate model |
| --- | --- | --- | --- |
| $\Lambda$ | 0.0080 | 0.0050 | 0.0050 |
| $c$ | 0.6300 | 0.6300 | 0.6300 |
| $\delta$ | 0.0001 | 0.0001 | 0.00009 |
| $\sigma$ | 0.9040 | 0.0552 | 0.0052 |
| $\beta$ | 1.0933 | 0.7564 | 0.6883 |
| $\gamma$ | 0.5888 | 0.5945 | 0.3970 |
| $\lambda$ | 0.5804 | 0.5631 | 0.3255 |
| $\theta$ | 0.1653 | 0.0421 | 0.0259 |
| Least square error | 3.9623e+2 | 3.2931e+2 | 3.2901e+2 |

**Table 4:** Estimated fractional orders.

| Model/fractional order | $\alpha_1$ | $\alpha_2$ | $\alpha_3$ | $\alpha_4$ |
| --- | --- | --- | --- | --- |
| Classical | 1 | 1 | 1 | 1 |
| Commensurate | 0.7894 | 0.7894 | 0.7894 | 0.7894 |
| Incommensurate | 0.8150 | 0.7683 | 0.8084 | 0.7834 |

**Figure 2:**
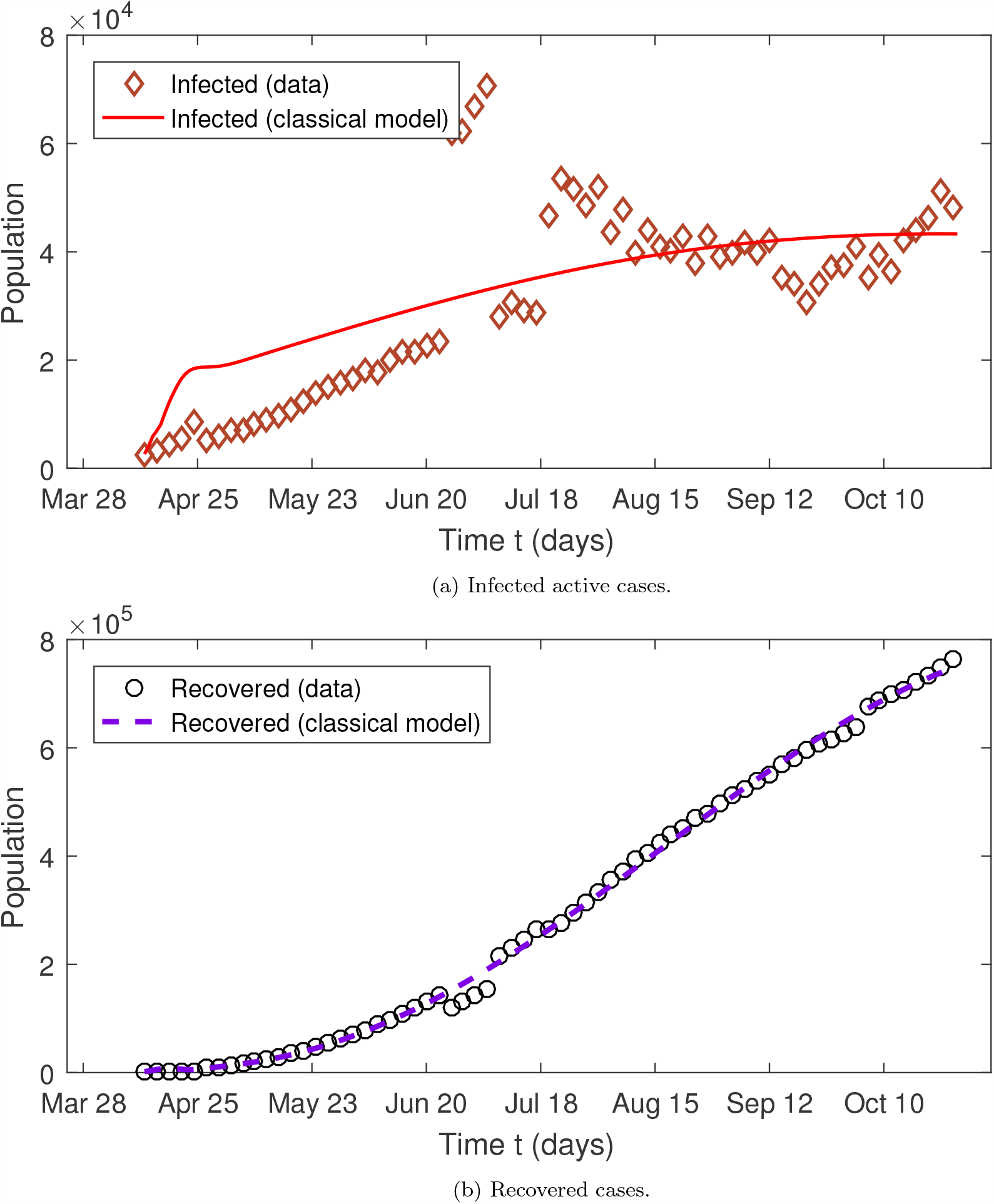
Graphical representation of the model fitting to the infected active cases and cumulative total of Recovered COVID-19 case in Mexico using the classical model (1). The period studied is from April 12 to October 28, 2020.

**Figure 3:**
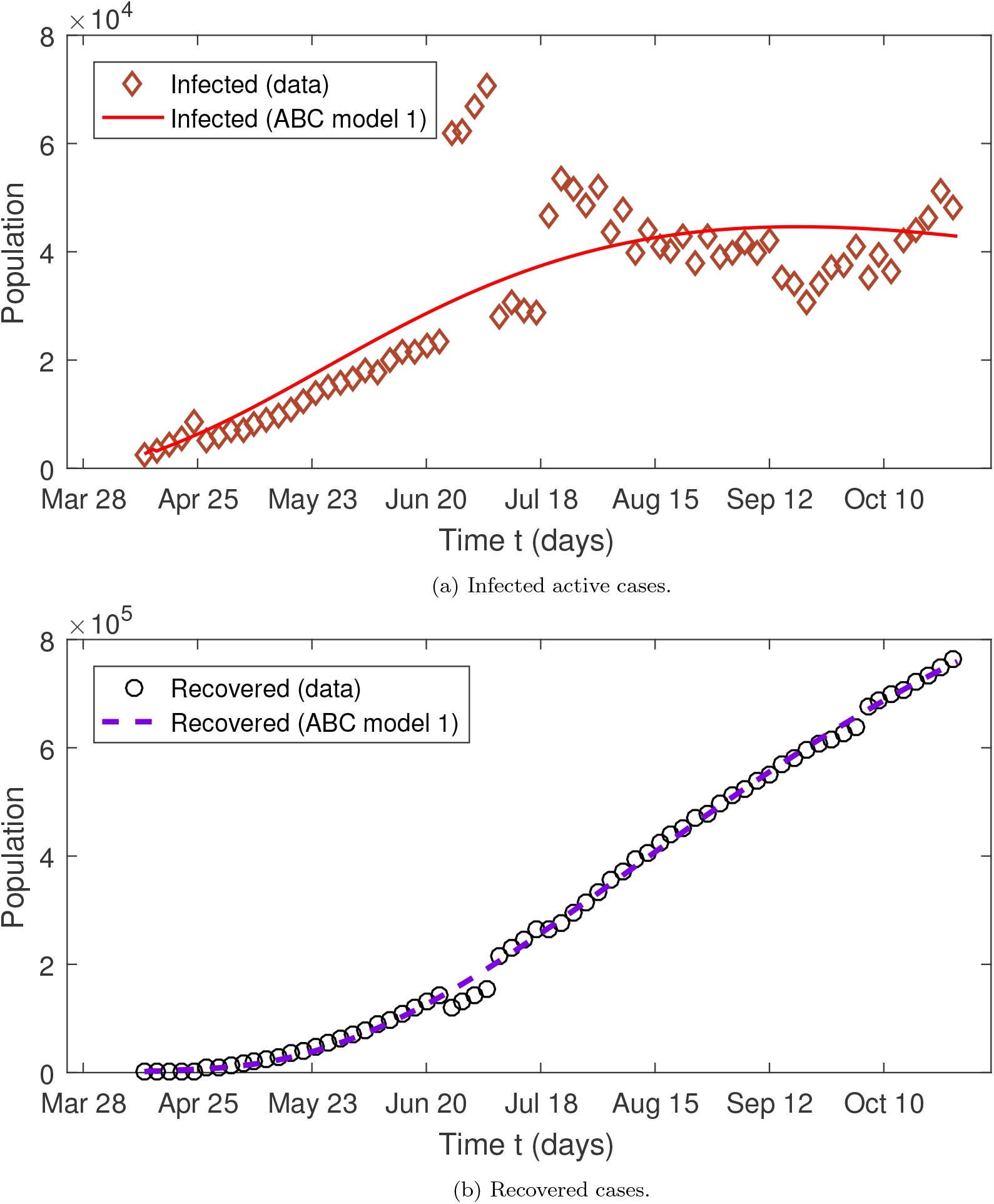
Graphical representation of the model fitting to the infected active cases and cumulative total of Recovered COVID-19 case in Mexico using the commensurate model (ABC model 1). The period studied is from April 12 to October 28, 2020.

**Figure 4:**
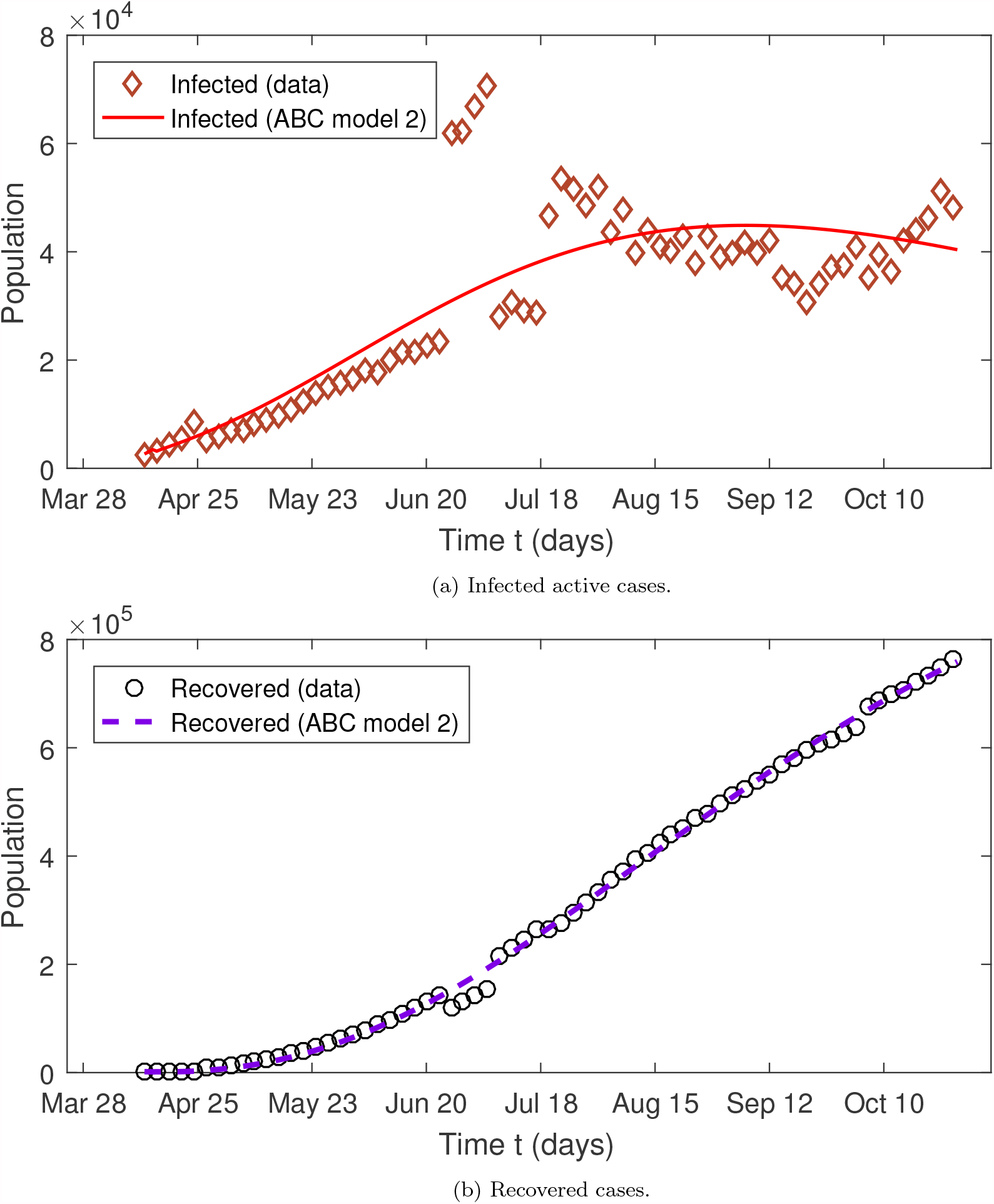
Graphical representation of the model fitting to the infected active cases and cumulative total of Recovered COVID-19 case in Mexico using the incommensurate model (ABC model 2). The period studied is from April 12 to October 28, 2020.

From Table 3 we notice that our models give a good approximation of COVID-19 real data in Mexico compared to the classical model and more better than the case in which the orders of the four model equations have the same fractional order. The error of least square for the commensurate and incommensurate models are respectively **3.2931e+2** and **3.2901e+2**, this means that the incommensurate model with ABC-fractional derivative gives a better fit of the real data compared to the commensurate model with ABC-fractional derivative. In general the ABC-fractional derivative gives results that are closer to the real data compared to the classical model.

### 7.2. Sensitivity analysis of model parameters with respect to ℛ_0_

When a virus spreads quickly it is necessary to look for methods to control the virus, that is, seek to increase or decrease some parameters which have an influence in the transmission of the virus. Sensitivity analysis is one way to determine the importance of each parameter for disease transmission.

Precisely, we use the definition of the sensitivity index which gives the significance of the variable associated with each parameter of the model to reduce the disease, the sensitivity index is defined as in [42].

#### Definition 9

*The sensitivity index of* ℛ_0_ *with respect to x, defined by*

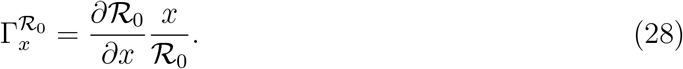

The sign of each index makes it possible to know whether the parameter increases (positive sign) or decreases (negative sign) the value of ℛ_0_,

Using the expression of ℛ_0_ given in (15) to calculate the sensitivity index for ℛ_0_ with respect to the parameters of the model, we will find

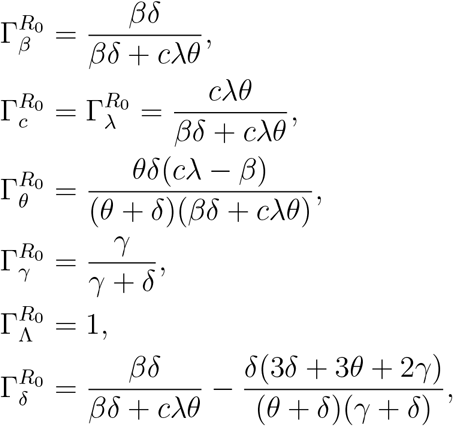

Note that ℛ_0_ does not depend on *σ* so 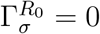.

From Table 5, we can see that 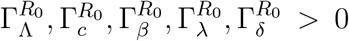 while 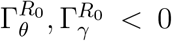 which means that an increment in Λ, *c, β, λ* and *δ* will cause ℛ_0_ to increase, while a decrement in the recovery rate of infected γ or the quarantine rate of susceptible *θ* will cause ℛ_0_ to decrease.

**Table 5:** Sensitivity analysis of *R*_0_, using the parameter values described in Table 1.

| Parameters | Classical model | Commensurate model | Incommensurate model |
| --- | --- | --- | --- |
| $\Lambda$ | +1.0000 | +1.0000 | +1.0000 |
| $c$ | +0.9982 | + 0.9950 | +0.9872 |
| $\delta$ | +0.0001 | +0.0002 | +0.0043 |
| $\sigma$ | 0 | 0 | 0 |
| $\beta$ | +0.0018 | + 0.0050 | + 0.0128 |
| $\gamma$ | -0.9998 | -0.9998 | -0.9997 |
| $\lambda$ | + 0.9982 | +0.9950 | +0.9872 |
| $\theta$ | -0.0012 | -0.0027 | -0.0089 |

### 7.3. Impact of fractional order

Based on numerical fractional Euler method derived in section (6), we present the graphical results of the proposed incommensurate fractional model (2) in Figures 5 and 6 to analyze the influence of fractional order. We have considered the initial values given in Table 2 and the estimated parameters given in Table 3.

**Figure 5:**
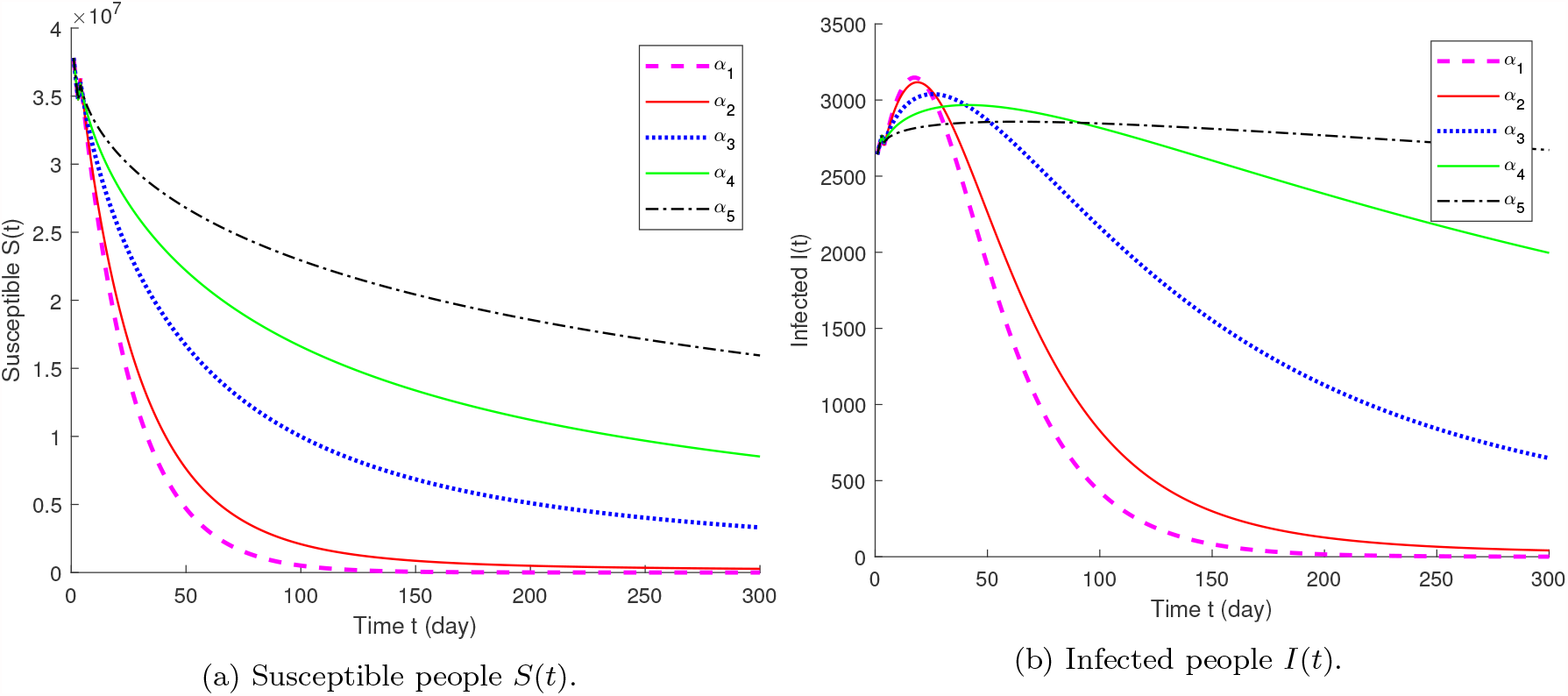
The representation of the numerical solution of the fractional model obtained for *S*(*t*) and *I*(*t*) for five arbitrary values of fractional multi-order. These results are obtained using the parameters presented in Table 3 except Λ and *δ* are chosen to be zero. We have chosen *α*_1_ = (1, 1, 1, 1), *α*_2_ = (0.95, 0.95, 0.95, 0.95), *α*_3_ = (0.8, 0.8, 0.8, 0.8), *α*_4_ = (0.7, 0.65, 0.6, 0.5) and *α*_5_ = (0.4, 0.2, 0.3, 0.1).

**Figure 6:**
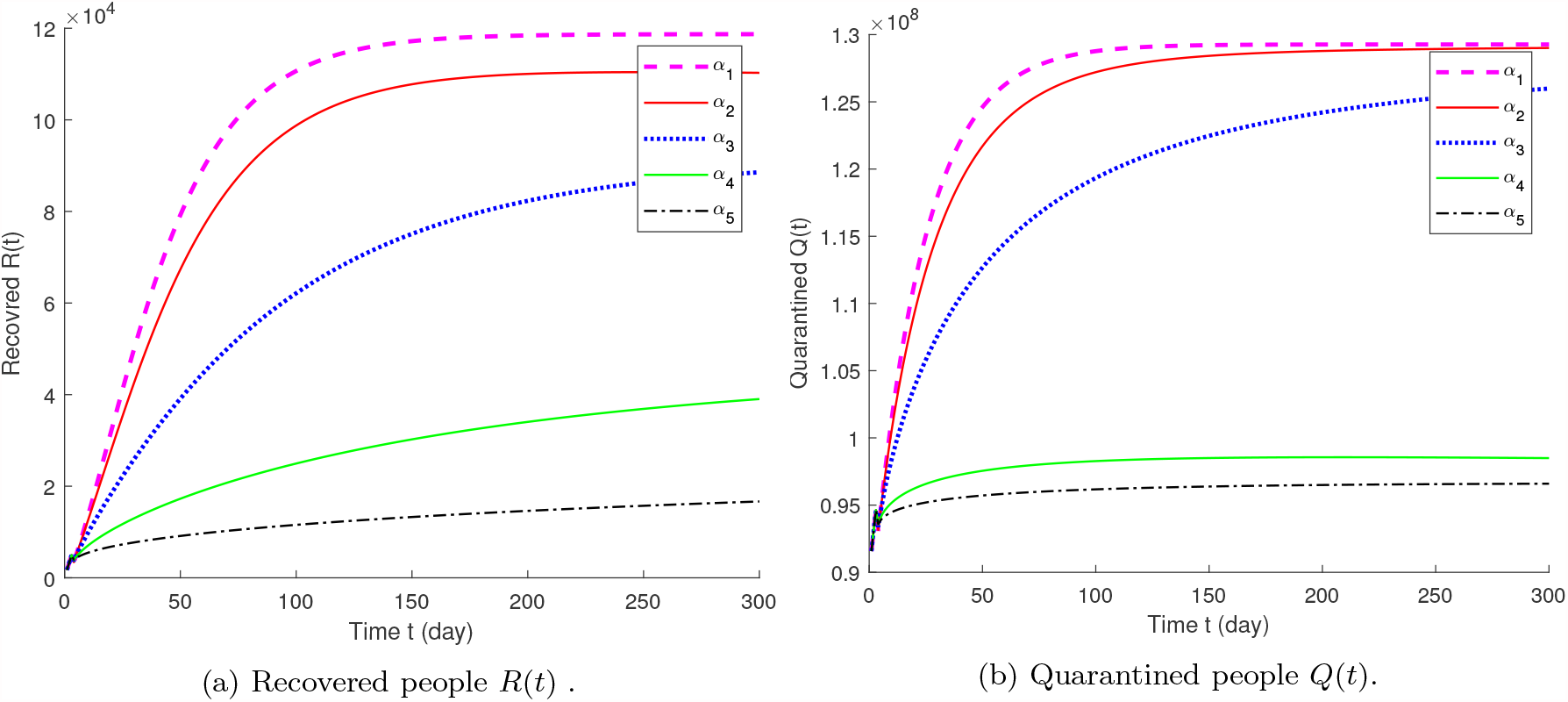
The representation of the numerical solution of the incomensurate fractional model obtained for *R*(*t*) and *Q*(*t*) for five arbitrary values of fractional multi-order. These results are obtained using the parameters presented in Table 3 except Λ and *δ* are chosen to be zero. We have chosen *α*_1_ = (1, 1, 1, 1), *α*_2_ = (0.95, 0.95, 0.95, 0.95), *α*_3_ = (0.8, 0.8, 0.8, 0.8), *α*_4_ = (0.7, 0.65, 0.6, 0.5) and *α*_5_ = (0.6, 0.5, 0.45, 0.4).

The obtained figures show the importance of choosing the fractional order in the 2019nCOV model, as well as choosing a different order in each equation. From Figures 5-6, we see that any slight change in the fractional orders gives a change in the dynamics of the infected people and this indicates the effectiveness of this type of derivation. It can also be used to predict the number of people infected with COVID-19 better than the classical model and the fractional commensurate model (see Table 3).

## 8. Conclusion

In this chapter we have studied an incommensurate fractional model of COVID-19 with Atangana-Baleanu fractional derivative, we have proved the existence together with the uniqueness of the solution using the fixed-point theorem. We have determined the equilibrium points of the proposed model and the expression of base reproduction number *R*_0_ as well as the local stability of each equilibrium point. The numerical fractional version of Euler method is used to approximate the solutions of the fractional model. We simulated the models with five different fractional order vector and we compared the results obtained. The model efficiency is shown by comparing the error values between the COVID-19 data in Mexico and the values predicted by the three models studied. The best fit for COVID-19 data in Mexico is established when using incommensurate model with Atangana-Baleanu operator. The Least Squares method is used to determine the best fit to real data and estimate fractional order. Finally, the chapter gives an example of the effectiveness of using the Atangana-Baleanu operators in real problems and the importance of incommensurate propriety in the fractional model.

## Data Availability

None

